# Systematic Observation of COVID-19 Mitigation (SOCOM): Assessing Face Covering and Distancing in Schools

**DOI:** 10.1101/2021.03.13.21253522

**Authors:** Ricky Camplain, Nanette V. Lopez, Dan M. Cooper, Thomas L. McKenzie, Kai Zheng, Shlomit Radom-Aizik

## Abstract

**Introduction:** During the COVID-19 pandemic, some K-12 schools resumed in-person classes with varying degrees of mitigation plans in the fall of 2020. Physical distancing and face coverings can minimize SARS-CoV-2 spread, the virus that causes COVID-19. However, no research has focused on mitigation strategy adherence during school days. Thus, we sought to develop a systematic observation protocol to capture COVID-19 mitigation strategy adherence in school environments: The Systematic Observation of COVID-19 Mitigation (SOCOM).

**Methods:** We extended previously validated and internationally used tools to develop the SOCOM training and implementation protocol to assess physical distancing and face covering behaviors. SOCOM was tested in diverse indoor and outdoor settings (classrooms, lunchrooms, physical education [PE], and recess) among diverse schools (elementary, secondary, and special needs).

**Results:** For the unique metrics of physical-distancing and face-covering behaviors, areas with more activity and a maximum of 10-15 students were ideal for accurately capturing data. Overall proportion of agreement was high for physical distancing (90.9%), face covering (88.6%), activity type (89.2%), and physical activity level (87.9%). Agreement was lowest during active recess, PE, and observation areas with ≥20 students.

**Conclusions:** Millions of children throughout the US are likely to return to school in the months ahead despite the current surge of COVID-19 cases. SOCOM is a relatively inexpensive tool that can be implemented by schools to determine mitigation strategy adherence and assess changes to protocols to help students return to school safely and slow the spread of COVID-19 and can be used for research purposes.

## INTRODUCTION

This work is to present a novel approach to assess the implementation of key mitigation procedures in children and adolescents in K-12 schools. The impact of the coronavirus disease 2019 (COVID-19), which is caused by the novel coronavirus, severe acute respiratory syndrome coronavirus 2 (SARS-Cov-2), has been widespread, with 21 million cases diagnosed and over 350,000 deaths in the U.S. as of January 4, 2020. When COVID-19 was declared a pandemic, schools were among the first operations to close to prevent community spread. However, in the fall of 2020, 56 million school-aged children (5-17 years of age) resumed education as some schools opened to in-person classes with varying degrees of public health mitigation plans.^1^ The extent to which children and adolescents comprise an asymptomatic population of SARS-CoV-2-infected individuals capable of spreading disease to school staff and family has yet to be resolved.^2^ Mitigation procedures can minimize SARS-CoV-2 outbreaks even in the close quarters of overnight summer camps.^3^

The K-12 school environment is particularly challenging with respect to the potential for viral transmission. Viral aerosolization can occur due to the general social and physically interactive nature of school-aged children, including during classroom learning, communal dining, recess, and PE classes. Comprehensive return-to-school plans emphasize adherence to accepted SARS-CoV-2 viral mitigation strategies procedures, including physical distancing (staying at least 6 feet from others who are not from the same household) and face cover wearing.^4-6^ Recognizing that these mitigation strategies represent unfamiliar and unnatural behaviors for K-12 students, attention has been focused on strategies to implement and operationalize mitigation protocols in schools. Little emphasis has been given to how to quantify adherence. Without metrics that gauge adherence to recommendations/rules, the short- and long-term effectiveness of SARS-CoV-2 transmission mitigation procedures cannot be ascertained.

Instruments are available for quantifying elements of behaviors that could be applied to assess the fidelity of COVID-19 mitigation. These include self-report,^7^ wearable technologies,^8-13^ and direct observation by trained personnel.^14-17^ Self-reports by children and adolescents of their daily living (e.g., physical activity or diet) are highly inaccurate.^14-17^ Wearable technologies such as accelerometers are increasingly used to gauge physical activity and sedentary behavior in children,^18,19^ but necessary technological advances in position and small-distance sensing are not available for widespread use and are not designed to assess physical distancing or face covering behaviors.^20-22^ Moreover, the use of cell phone-or GPS-monitoring in school-aged children and adolescents would raise questions of health data privacy,^23^ embodied in FERPA and HIPAA regulations.

Long-existing, robust, and widely used direct observation approaches developed to assess levels of physical activity in groups of children, adolescents, and adults in a variety of real-world environments were readily adapted to measure COVID-19 mitigation strategies in K-12 schools. The original System for Observing Play and Leisure Activity in Youth (SOPLAY), designed for physical activity settings in schools, and the System for Observing Play and Recreation in Communities (SOPARC), designed to include playgrounds and parks, were designed to obtain direct information on physical activity in open spaces.^16,24^ SOPLAY and SOPARC are based on momentary time sampling techniques in which systematic and periodic scans of individuals and contextual factors in physical activity environments are made and they have been adapted for multiple settings.^17^ Given SOPARC’s proven ability to obtain reliable observational data, we sought to test a new strategy of using systematic observation to capture COVID-19 mitigation strategy adherence, including physical distancing and face coverings, among grade-school-aged children in diverse school settings: The Systematic Observation of COVID-19 Mitigation (SOCOM).

## MATERIALS AND METHODS

### Study Setting

Four schools in Orange County, California, were recruited to participate in a Safe School Restart study to begin essential research on COVID-19 transmission in children and adolescents as K-12 schools reopened across the U.S. The aim of Safe School Restart was to determine how effectively state and regional guidelines slowed viral transmission as schools restarted. A hypothesis of the study is that mitigation protocol fidelity will be related to the relative socio-economic condition of a school. Private schools across the country have been able to expend the resources necessary to develop and implement effective mitigation strategies. Public schools, particularly Title 1 public schools, have had greater difficulty in accessing necessary resources.^25,26^ Consequently, our study includes four representative schools in Orange County, California:

1. Private K-12 school serving predominantly middle- and upper-middle socioeconomic status (SES) students
2. K-6 public school serving predominantly Latino, lower SES students
3. K-8 public charter school located in a predominantly Latino, lower SES neighborhood
4. K-6 public charter school serving predominantly children with special needs, including down syndrome, autism spectrum disorders, etc.

SOCOM was designed to observe four distinct times during the school day: classroom learning (non-physical activity courses taught in a classroom), active recess (a regularly scheduled period in the school day for physical activity and play that is monitored by trained staff or volunteers), PE class (instruction in physical exercise and games in school), and communal dining (lunch) time.

### Partnerships with Schools

Schools were selected for Safe School Restart to reflect the wide range of SES, ethnic diversity, and school layouts and facilities that exist within Orange County, California, allowing to determine the ability of observers to implement SOCOM in different settings (Table 1). Previous relationships with the schools facilitated approvals by schools and school districts. Great care was taken to plan with school staff, so elements of the study were clear, acceptable, and followed the policies taken by the schools in response to the COVID-19 pandemic.

**Table 1.**
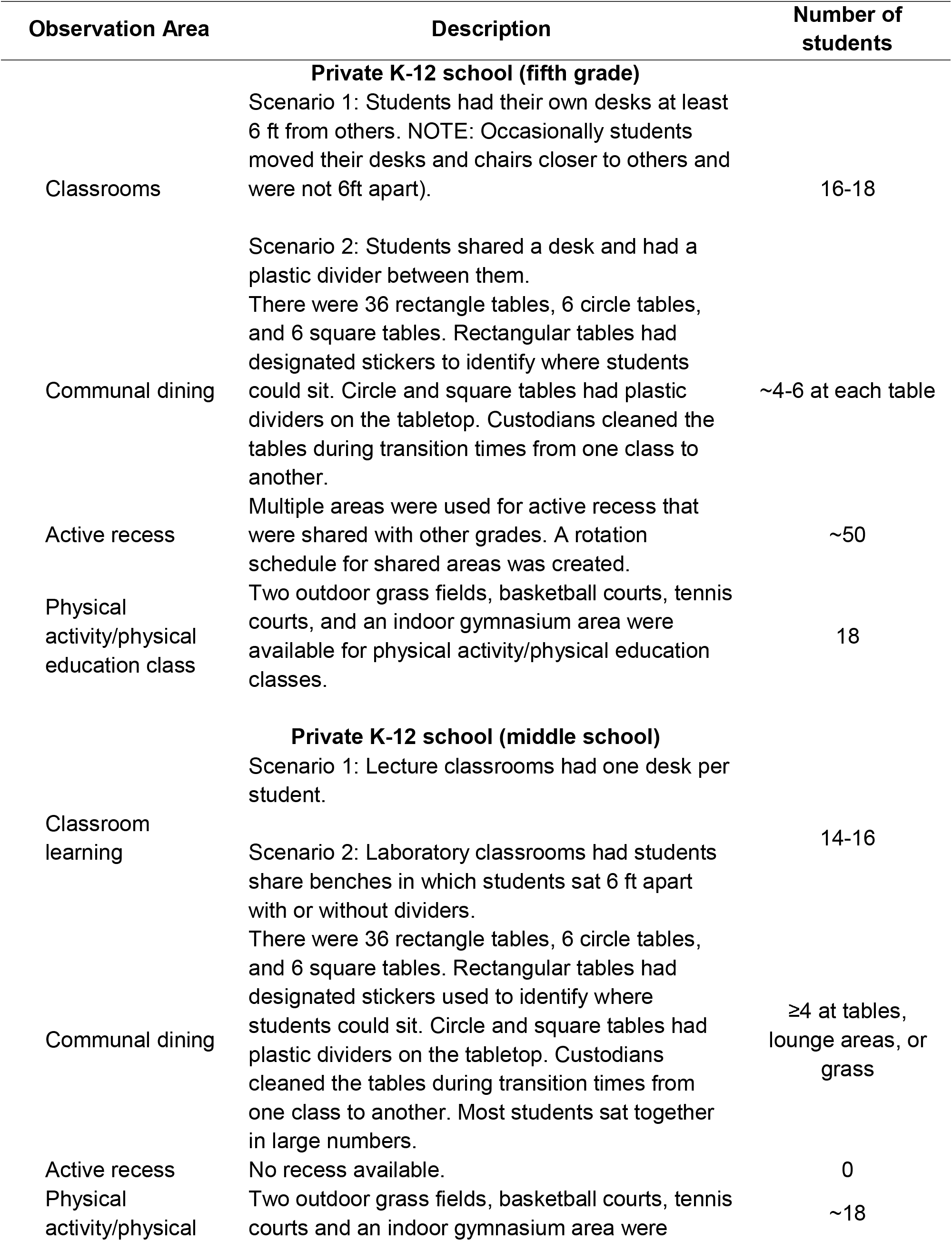
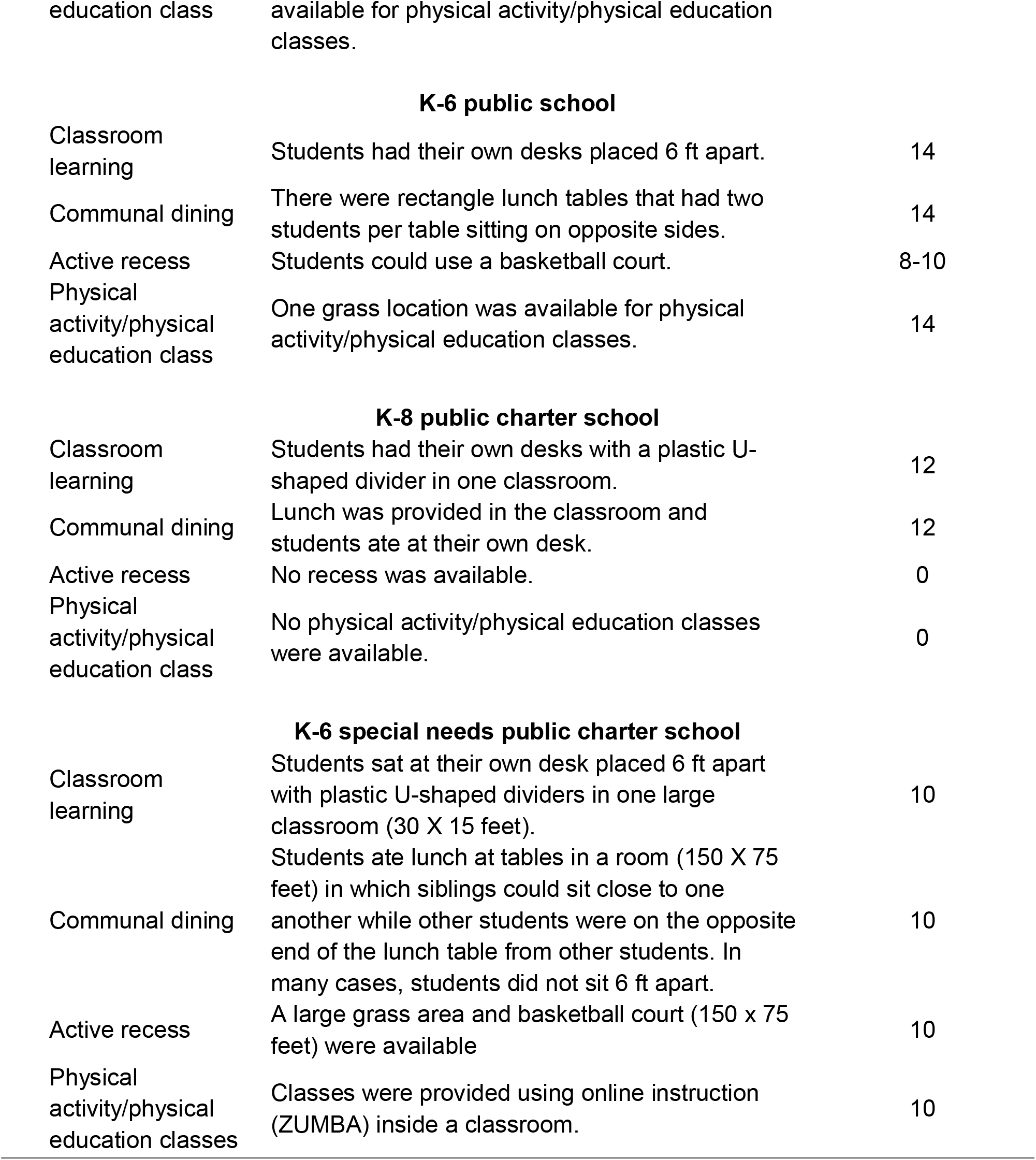
Characteristics Within Observed Schools.

### SOCOM Development

In the summer of 2020, our team, in collaboration with physical activity and SOPARC experts, developed the SOCOM protocol to focus on assessing COVID-19 mitigation procedures (physical distancing and face covering behaviors). Paper and electronic observation forms were developed to capture data (Figures 1 and 2). The custom electronic data capture system optimized for mobile devices was developed by the Center for Biomedical Informatics, UC Irvine Institute for Clinical and Translational Science. The platform leverages the Bootstrap responsive user interface framework and was programmed in ASP.Net C#. In addition to the SOPARC-based data fields that research coordinators can enter manually based on their field observations, the system is also capable of obtaining local weather information automatically through an application programming interface (API) provided by openweathermap.org. All data captured are stored in a HIPAA-compatible environment hosted at the Enterprise Data Center of the UCI Health.

**Figure 1.** Sample Systematic Observation of COVID-19 Mitigation (SOCOM) Electronic Data Collection Form.

**Figure 2.** Sample Systematic Observation of COVID-19 Mitigation (SOCOM) Paper Data Collection Form.

### Measures

Primary measures included physical distancing, face covering behavior, physical activity levels, and activity type (Table 2). In addition, observed individuals were categorized into sex/age groups (female students, male students, female adults, and male adults). Data were recorded to identify specific observation target area, date, grades in attendance, time of observation, and weather conditions.

**Table 2.** Systematic Observation of COVID-19 Mitigation (SOCOM) Central Measures.

| Variable | Definition |
| --- | --- |
| Physical distancing | 1. *Physically distanced* – observed individual was not within six feet of another person<br>2. *Not physically distanced* – individual being observed was within six feet of others<br>3. *Unknown* – observer could not determine if observed individual was within six feet of others |
| Face covering behavior | 1. *On* – individual had both his/her nose and mouth fully covered<br>2. *Partial* – individual had face covering on but either his/her nose or mouth was not covered<br>3. *Not on* – individual had no face covering on, but one was visible (e.g., mask in hand or hanging from ear)<br>4. *None* – individual had no visible face covering anywhere on body |
| Physical activity levels | 1. *Sedentary* – individual was lying down, sitting, or standing in place<br>2. *Walking* – individual was engaged in walking at normal pace<br>3. *Vigorous* – individual was engaged in an activity more vigorous than an ordinary walk (e.g., causing increased heart rate, such as when jogging, swinging, doing cartwheels). |
| Activity type | 1. *Interactive* – individual was engaged in an interactive activity where he/she was using a shared piece of equipment or touched another person<br>2. *Individual* – individual was engaged in an independent activity with no shared equipment |

### Training

In preparation for SOCOM, observers studied the written protocol and then attended a four-hour workshop. Original written and video SOPARC training materials were used during the workshop (https://activelivingresearch.org/soparc-system-observing-play-and-recreation-communities). Training included reviewing definitions and coding conventions, differentiating among codes, coding practice, and orientation to the observation tools. Observer preparation also included target area mapping strategies. The target areas (i.e., pre-determined observation locations) were diverse and varied by school. Coding conventions and how to differentiate between the various physical activity levels included video lectures and practice with feedback from videotaped samples through original SOPARC training. Distinguishing between face coverings, physical distancing, and activity type definitions was taught using photographs and videos. Observers practiced coding and received feedback on their scoring before engaging in field practice.

### Observation Preparation

In addition to observer training, the research team obtained maps of the school grounds from Google Maps and/or school administrators. These maps were used to precisely identify outdoor target areas, spaces in which activities may occur and are small enough to accurately count people using them. The size, location, and boundaries for both indoor and outdoor target areas were identified and the sequential order for observing them established.

As COVID-19 recommendations and restrictions varied among schools and time periods, visiting schools was imperative to finalizing school-specific observation protocols and visitation days. Administrative-level school officials (e.g., principals) escorted observers around a school and its grounds to note the general layout and major features of the facilities and to specifically identify potential target areas for observation.

Finally, before official data collection began, all observers attended a field-based training at one of the schools using the developed protocol and data collection form to practice using the protocol and form. Observers used the schedule of observations similar to that for official observations, but were able to discuss protocols and category classifications.

### Recording Procedures

To establish reliability, during visits to a target area, two to three observers simultaneously completed the observation form either using the paper form or individual cell phones (Figure 1 and 2), and timestamps and weather information were uploaded automatically into the electronic form.

After recording the characteristics of a target area, the observers completed scans of the area by making independent “visual sweeps” from left to right, in which two or three observers scan simultaneously at the same pace. Separate scans were conducted for each sex/age group (female students, male students, female adults, and male adults), and each characteristic (physical distancing, face covering, physical activity, and activity type) for a maximum of 16 scans per target area. For example, during the first scan for female students, observers recorded whether each was physically distanced, not physically distanced, or unknown. During the second scan, observers recorded data for each female student’s face covering (e.g., on, partial, not on, or no mask). For PE class and active recesses only, a third scan was performed to record the physical activity level of each female student as sedentary, walking, or vigorous. Finally, for female students a scan was made to categorize each as being interactive or individual. This scanning procedure was then completed for male students, female adults, and male adults in the target area (if any). After this, observers moved to the next specified target area. Occasionally (e.g., unusual large numbers of people in the area or full view of the area being obstructed), target areas were subdivided into smaller sub-areas so more accurate measures could be obtained. Data from these sub-areas were later summed to provide an overall measure for each target area.

During groups of scans, observers were also able to take qualitative notes on contextual information. When needed, observers used the paper observation form and later entered the information into the electronic data form.

### Planned Data Analysis

For physical distancing scenario, face covering behavior, physical activity level, and activity type, counts will be tallied for those engaged in each group in each school and observation area to obtain a summary score for female students, male students, female adults, and male adults. A proportion of individuals in each physical distancing scenario, face covering behavior, physical activity level, and activity type will be calculated by age/sex group.

## RESULTS

### Observations

During a school visit, the time for observations varied by the number of target areas and the number of people present. Observation length for target areas ranged from about two (classrooms) to 15 minutes (dining rooms). Communal areas generally had more people and more target areas, so more scans were required. Additionally, when more people were present (>20), some observers preferred the paper observation form compared to the electronic form as they could easily tally students and take notes in the margins of the form. Thus, for physical distancing and face covering, areas with more activity and target areas, observation areas with a maximum of 10-15 students was ideal for accurately and confidently capturing data. Among target areas with primarily sedentary people (e.g., classrooms), observers felt more comfortable with a maximum of 15-20 students.

The presence of observers being adjacent to target areas, especially indoor ones, may have slightly influenced staff and student behavior. Schools had restrictions on visitors due to COVID-19 protocols [e.g., number of visitors allowed in a room and teachers sometimes inquired about why observers were there (during their first visit)]. Meanwhile, when students were more physically active (e.g., PE classes) observers typically went unnoticed.

### Reliability and Validity

Reliability data for physical activity codes, physical distancing, and face covering behaviors among students were collected during all observation periods using two or three assessors who made simultaneous, independent observations. Data from 166 scans were used in the reliability of physical distancing, face covering behaviors, and activity type analyses, and the overall proportion of agreement was calculated for each variable (face covering, 88.6%; physical distancing, 90.9%; and activity type, 89.2%).

Reliability for physical activity level was calculated for 34 observations (collected during active recess and PE classes) and the proportion of agreement was 87.9%. When assessed by observation area (classroom learning, communal dining, active recess, and PE classes), agreement was lowest during active recess and PE classes (79.7% to 99.1%). Agreement was lower when there were ≥20 students present (80.2%) compared to <20 students (90.2%).

Original SOPARC activity codes were used in other observation systems including Behaviors of Eating and Activity for Children’s Health: Evaluation System (BEACHES),^27^ the System for Observing Fitness Instruction Time (SOFIT),^28^ and SOPLAY.^16^ Construct validity of activity codes has been established through heart rate monitoring among children 4-18 years old^27,29^ and with accelerometers in schools.^30^ We did not determine validity of the physical activity codes in this study.

## DISCUSSION

Systematic observation has been widely used in collecting information on children’s and adolescents’ behaviors.^16,17,24,27^ Therefore, we sought to test a new strategy of using systematic observation to capture COVID-19 mitigation strategy adherence, including physical distancing and face coverings, among grade-school-aged children in the school environment. The initial experience with SOCOM indicates that it may serve as a reliable and useful tool in assessing COVID-19 mitigation procedures in schools.

Interrater reliability profiles for SOCOM, including face covering, physical distancing adherence, and physical activity were comparable to these metrics in SOPLAY and SOPARC.^24^ After and while the team completed observations, the research team discussed the effectiveness of the SOCOM protocol, observation form, and experience. In general, heuristic assessments of the SOCOM protocol and observation forms were positive with certain limitations in using the electronic form, including the need for access to the internet on the devices used. SOCOM worked well, even in the somewhat constrained setting of school environments during a pandemic.

Due to the complex, novel nature of capturing COVID-19 mitigation strategy adherence, training and observation preparation were extremely important. Training materials for SOCOM were obtained from existing valid and reliable methods from the original SOPARC tool. Although previous training materials focused mainly on capturing physical activity levels, we found that training to observe physical distancing and face covering behaviors was readily adaptable. Field-based observational training at the schools using the protocol in real-time were imperative. Individual schools had different layouts for classrooms, meals, recesses, and physical activity facilities (e.g., playgrounds). Consequently, our training included an initial, separate visit to each school to familiarize the observers with the features of the school’s physical environment and to review details of the school’s COVID-19 schedule, rules, and restrictions. This permitted the development of appropriate weekly and daily observation schedules for each school and was welcomed by school leadership and staff. Visits were necessary for planning the positioning observers in areas where they could have a clear view and to minimize interactions with students and staff. However, a limitation of SOCOM is the potential for observation bias as individuals may be inclined to be on their best behavior while under observation.

With high reliability, ease of use, inexpensive minimal training, and the ability to modify the tool as needed, SOCOM could be used by other researchers, school administration, and school staff to help schools reopen successfully. The tool can be applied by schools to monitor compliance, adjust mitigation strategy messaging, and contribute to informed school policies. Furthermore, the tool can be adjusted to use in other school settings, such as music classes, lab settings, teachers lounges, and other parts of the school. Although SOCOM may be adapted for use beyond schools, we did not observe those settings.

## CONCLUSIONS

Despite the current surge of COVID-19 cases, millions of children and staff will be in schools under various conditions around the world. Despite the inauguration of COVID-19 vaccinations, the vaccination of enough people to approach herd immunity is a long way off. At the time of this writing, a vaccine has not been approved to be used among children <16 year old and no child under the age of 12 years has been enrolled in any safety or efficacy vaccine trials, thus the potential for widespread COVID-19 vaccinations in school-aged children to diminish the need for mitigation procedures in schools is many months away. There is mounting evidence of the damage done to K-12 students, particularly low-income and minority, by continued school closures,^31,32^ and it is likely that increasing numbers of schools will reopen by the beginning of the 2021 winter semester. Quantifying the success of SARS-CoV-2 transmission mitigation in school settings is likely to be useful for the foreseeable future. Standardized methods of measuring the fidelity of mitigation procedures will likely aid in identifying the most effective ways to minimize SARS-CoV-2 viral transmission. SOCOM is a relatively inexpensive tool that can be implemented by schools in various settings, including the school day, after school programs, and school sports and competitions, to determine mitigation strategy adherence to help students return to school safely and slow the spread of COVID-19.

## Data Availability

Data are available upon request and agreement of the coauthors of the manuscript.

## ACKNOWLEDGEMENTS

The research team would like to thank Cherryl Nugas, data collection coordinator and observer, and Hoang Pham, Rula Ballat, and Rebecca Mai Huynh, observers, from the Pediatric Exercise and Genomics Research Center for their dedication to the project. We would additionally like to thank James F. Sallis, PhD, and Deborah A. Cohen, MD, for thoughtful insight and guidance during the development phases of the study.

The project was funded by the University of California Irvine (UC Irvine) Clinical and Translational Science Award (NCATS UL1; TR001414), the Orange County Health Care Agency, UC Irvine COVID-19 Research Fund, and the Northern Arizona University Southwest Health Equity Research Collaborative (NIH/NIMHD U54; U54MD012388).

